# Neutralization breadth of SARS CoV-2 viral variants following primary series and booster SARS CoV-2 vaccines in patients with cancer

**DOI:** 10.1101/2021.11.10.21265988

**Authors:** Vivek Naranbhai, Kerri J St. Denis, Evan C Lam, Onosereme Ofoman, Wilfredo-Garcia Beltran, Cristhian Berrios, Atul K. Bhan, Justin F. Gainor, Alejandro B. Balazs, A. John Iafrate, on behalf of the CANVAX team

**Author notes:** **Corresponding author:** A. John Iafrate, Massachusetts General Hospital Cancer Center, 55 Fruit Street, Boston, MA, 02114.

## Abstract

Patients with cancer are more likely to have impaired immune responses to SARS CoV-2 vaccines. We studied the breadth of responses against SARS CoV-2 variants followingly primary vaccination in 178 patients with a variety of tumor types, and after booster doses in a subset. Neutralization of alpha, beta, gamma and delta SARS-CoV-2 variants was impaired relative to wildtype (Wuhan), regardless of vaccine type. Regardless of viral variant, mRNA1273 was the most immunogenic, followed by BNT162b2 and then Ad26.COV2.S. Neutralization of more variants (breadth) was associated with higher magnitude of wildtype neutralization, and increase with time since vaccination; increased age associated with lower breadth. Anti-spike binding antibody concentrations were a good surrogate for breadth (PPV=90% at >1000U/ml). Booster SARS-CoV-2 vaccines conferred enhanced breadth. These data suggest that achieving a high antibody titer is desirable to achieve broad neutralization; a single booster dose with current vaccines increases breadth of responses against variants.

## Introduction

Patients with cancer are at increased risk of severe disease and/or death from SARS CoV-2 infection (Kuderer et al., 2020) and vaccination against SARS-CoV-2 is a cornerstone of prevention. The magnitude of SARS-CoV-2 spike or receptor-binding (RBD) antibodies and neutralization titer against wildtype SARS-CoV-2 are robust correlates of vaccine-mediated protection (Earle et al., 2021; Khoury et al., 2021) but are not sufficiently validated for use clinically. In the Cancer, COVID and Vaccination study (CANVAX) of more than 750 patients with cancer, we observed lower humoral immune responses than in non-cancer controls (Naranbhai et al., JCO In press), consistent with findings from other cohorts (Addeo et al., 2021; Bird et al., 2021; Thakkar et al., 2021). In both non-cancer controls and patients with cancer, the magnitude of response was strongly associated with prior SARS CoV-2 infection and vaccine type: mRNA1273 was the most immunogenic followed by BNT162b2. Both mRNA vaccines were markedly more immunogenic than Ad26.COV2.S. Finally, booster vaccines were able to overcome poor responses in CANVAX and other studies(Greenberger et al., 2021).

Evolution of SARS CoV-2 variants with mutations that confer higher transmissibility or evasion of immune responses pose an ongoing threat, as exemplified by the rapid rise of the delta variant to global dominance during 2021. We, and others, have previously observed marked variation of *in vitro* neutralization of viral variants in healthy individuals (Tada et al., 2021; Wilfredo Garcia-Beltran et al., 2021). There are few robust data regarding the degree of protection against each variant following different vaccines in immunocompromised patients, but the frequency of breakthrough infection resulting in hospitalization appears to be markedly higher for immunocompromised patients than in the general population, highlighting the impact of lower immunogenicity and higher risk of severe disease (Hippisley-Cox et al., 2021). Based on these and other data, additional ‘booster’ vaccine doses have been recommended for immunocompromised patients in many developed countries. Whilst these vaccine increase the magnitude of response(Greenberger et al., 2021), whether homologous (i.e. wildtype strain based) ‘booster’ doses enhance the breadth of protection against variants is uncertain (Cho et al., 2021).

We studied the magnitude and breadth of neutralization of SARS CoV-2 variants following the primary series, and after booster doses of vaccination in patients with cancer who received one of the SARS-CoV-2 Federal Drug Administration (FDA) Emergency Use Authorized (EUA) vaccines in the United States.

## Results

The CANVAX study is an ongoing prospective cohort study of SARS CoV-2 vaccines in patients with cancer. For this report, we selected 178 participants of CANVAX without prior SARS CoV-2 infection who were sampled >=14 days after vaccination stratifying by vaccine type (58 mRNA1273, 60 BNT162b2 and 60 Ad26.COV2S). The baseline participant characteristics known to affect immunogenicity are shown according to vaccine type in Table 1, and recapitulate those of the overall CANVAX study: Ad26.COV2.S recipients were slightly older but the sex, cancer type, and therapy types were similar between groups.

**Table 1:**
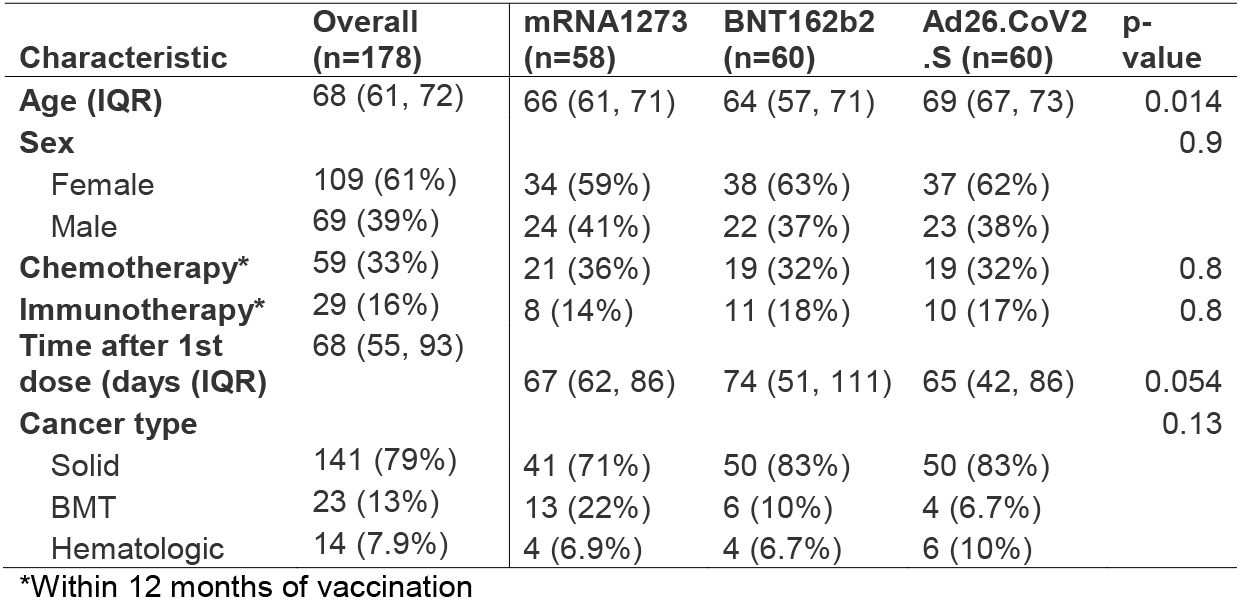
Baseline characteristics of participants in this study.

| Characteristic | Overall<br>(n=178) | mRNA1273<br>(n=58) | BNT162b2<br>(n=60) | Ad26.CoV2<br>.S (n=60) | p-value |
| --- | --- | --- | --- | --- | --- |
| **Age (IQR)** | 68 (61, 72) | 66 (61, 71) | 64 (57, 71) | 69 (67, 73) | 0.014 |
| **Sex** |  |  |  |  | 0.9 |
| Female | 109 (61%) | 34 (59%) | 38 (63%) | 37 (62%) |  |
| Male | 69 (39%) | 24 (41%) | 22 (37%) | 23 (38%) |  |
| **Chemotherapy\*** | 59 (33%) | 21 (36%) | 19 (32%) | 19 (32%) | 0.8 |
| **Immunotherapy\*** | 29 (16%) | 8 (14%) | 11 (18%) | 10 (17%) | 0.8 |
| **Time after 1st**<br>**dose (days (IQR))** | 68 (55, 93) | 67 (62, 86) | 74 (51, 111) | 65 (42, 86) | 0.054 |
| **Cancer type** |  |  |  |  | 0.13 |
| Solid | 141 (79%) | 41 (71%) | 50 (83%) | 50 (83%) |  |
| BMT | 23 (13%) | 13 (22%) | 6 (10%) | 4 (6.7%) |  |
| Hematologic | 14 (7.9%) | 4 (6.9%) | 4 (6.7%) | 6 (10%) |  |

\*Within 12 months of vaccination

### Neutralization of alpha, beta, gamma and delta SARS-CoV-2 variants is impaired in vaccine recipients

We assessed *in vitro* neutralization of wildtype SARS -CoV-2 (Wuhan strain) and four viral variants alpha, beta, delta and gamma using an extensively validated high-throughput pseudovirus neutralization assay (Garcia-Beltran et al., 2021a). These variants represent recent waves of the pandemic, and harbor both shared and distinct mutations (Supplementary Table 1) that are targeted by immune responses induced by vaccination with current vaccines, which all encode wildtype SARS CoV-2 spike protein. We quantified the serum pseudovirus neutralization titer (pNT50) associated with 50% reduction in viral entry into ACE2 expressing 293T-cells.

Consistent with the overall CANVAX population and other studies (Naranbhai et al., 2021; Tada et al., 2021), neutralization of wildtype SARS-CoV-2 was highest for mRNA1273 recipients, followed by BNT162b2 and lowest amongst Ad26.COV2.S recipients. Adjusting for covariates, neutralization was lower among BNT162b2 recipients than mRNA1273 for alpha, gamma and delta variants (Figure 1 and Supplementary Table 2).

**Figure 1:**
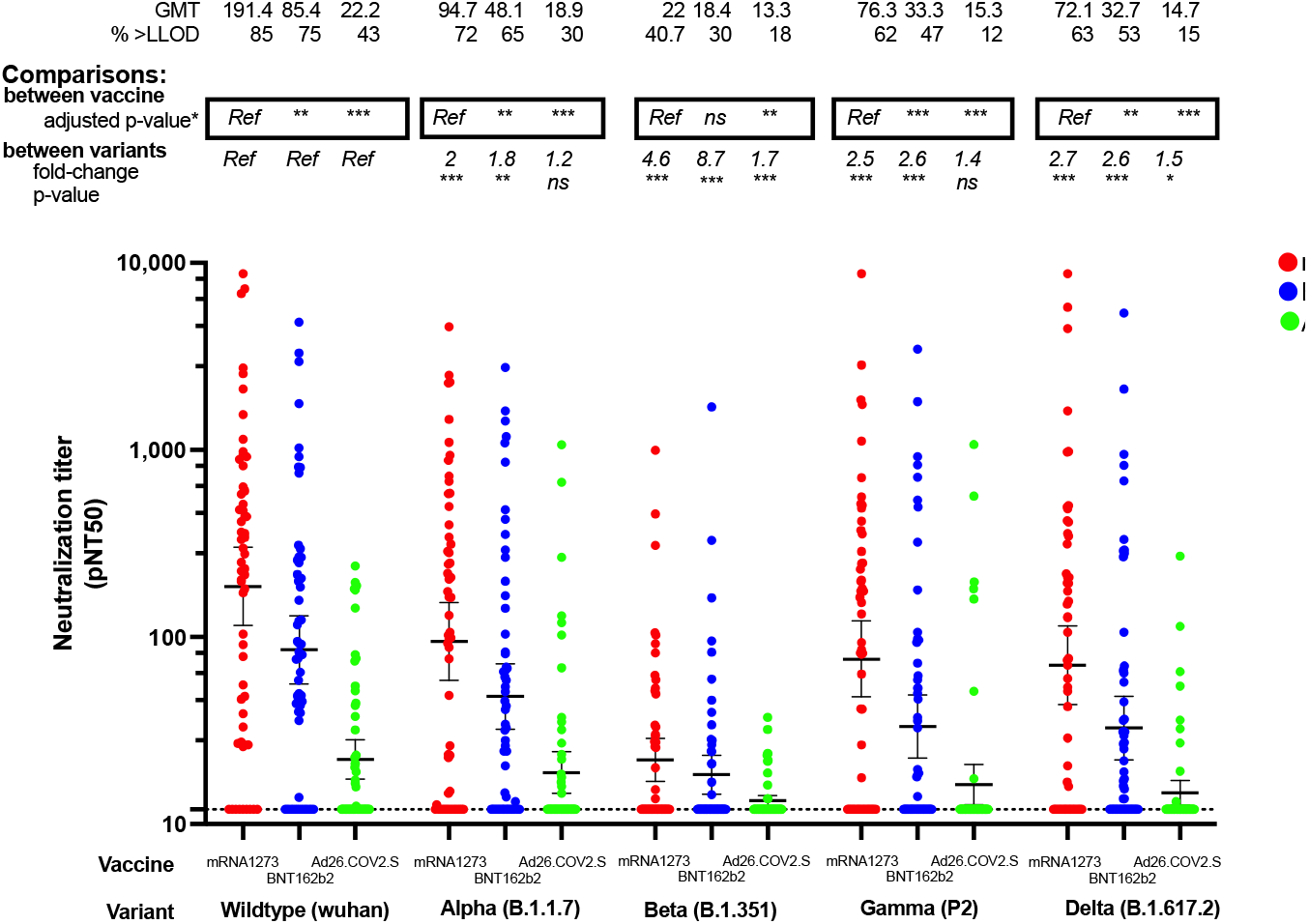
Neutralization of SARS CoV-2 variants following vaccination with mRNA1273 (n=58), BNT162b2 (n=60) or Ad26.COV2.S (n=60) in patients with cancer. The y-axis shows pseudovirus neutralization titer 50 (pNT50, defined as the titer at which the serum achieves 50% neutralization of SARS-CoV-2 wildtype pseudovirus entry into ACE2 expressing 293T cells).. Briefly, lentiviral particles encoding both luciferase and ZsGreen reporter genes were pseudotyped with the SARS-CoV-2 spike protein from the strain indicated (see Supplementary Table 1 for sequences) and produced in 293T cells, titered using ZsGreen expression by flow cytometry and used in an automated neutralization assay with 50–250 infectious units of pseudovirus co-incubated with three-fold serial dilutions of serum for 1 h. Neutralization was determined on 293T-ACE2 cells. A horizontal dotted line is shown at a pNT50 titer of 12 which is the lower limit of detection (LLOD) of this assay. The geometric mean titer, proportion positive (at a threshold of 1:12). Statistical comparison of neutralization titers against each strain between recipients of different vaccines is details in Supplementary Table 2 and denoted by ^*^ on the graph where p-value shows are adjusted for covariates previously shown to be associated with wildtype virus neutralization (REF) namely age, chemotherapy, immunotherapy, timing after vaccination, cancer type. Comparison of neutralization titers for recipients of each vaccine type, against different strains is shown as the fold change in neutralization, and corresponding p-value (based on a dunnet’s test conducted in GraphPad Prism v9.0).

We observed marked and significantly neutralization of alpha, beta, delta and gamma variants for most vaccine groups (Figure 1). The differences were most striking for the beta variant, where fewer than half of all evaluated donors (41% of mRNA1273, 30% of BNT162b and 18% of Ad26.COV2.S recipients) had neutralization measurable above the assay limit of detection. Few patients with cancer in this study had measurable neutralization against wildtype SARS-CoV-2 and the four variants tested following receipt of Ad26.COV2.S (43% against wildtype, 30% against alpha, 18% against beta, 12% against gamma and 15% against delta).

### Neutralization breadth is associated with wildtype neutralization titer, time since vaccination and is reduced with age

Next, we sought to identify correlates of vaccine breadth. In multivariate regression, the magnitude of neutralization of wildtype SARS-CoV-2 was the strongest correlate of breadth (effect estimate 1.4 additional variants neutralized per log_10_ increase in neutralization titer, 95% CI 1.1:1.6, adjusted p<0.001; Table 2). Vaccine type did not independently associate with breadth of neutralization (p>0.1 for each vaccine). Older age was associated with narrower breadth of response (effect estimate per 5 year increase in age -0.09, 95% CI -0.17:-0.01, adjusted p=0.029). Breadth of response tended to increase with time after vaccination, even adjusting for the expected initial increase and later waning in the magnitude of response (effect estimate per week after vaccination 0.04, 95% CI 0:0.08, adjusted p=0.061).

**Table 2:**
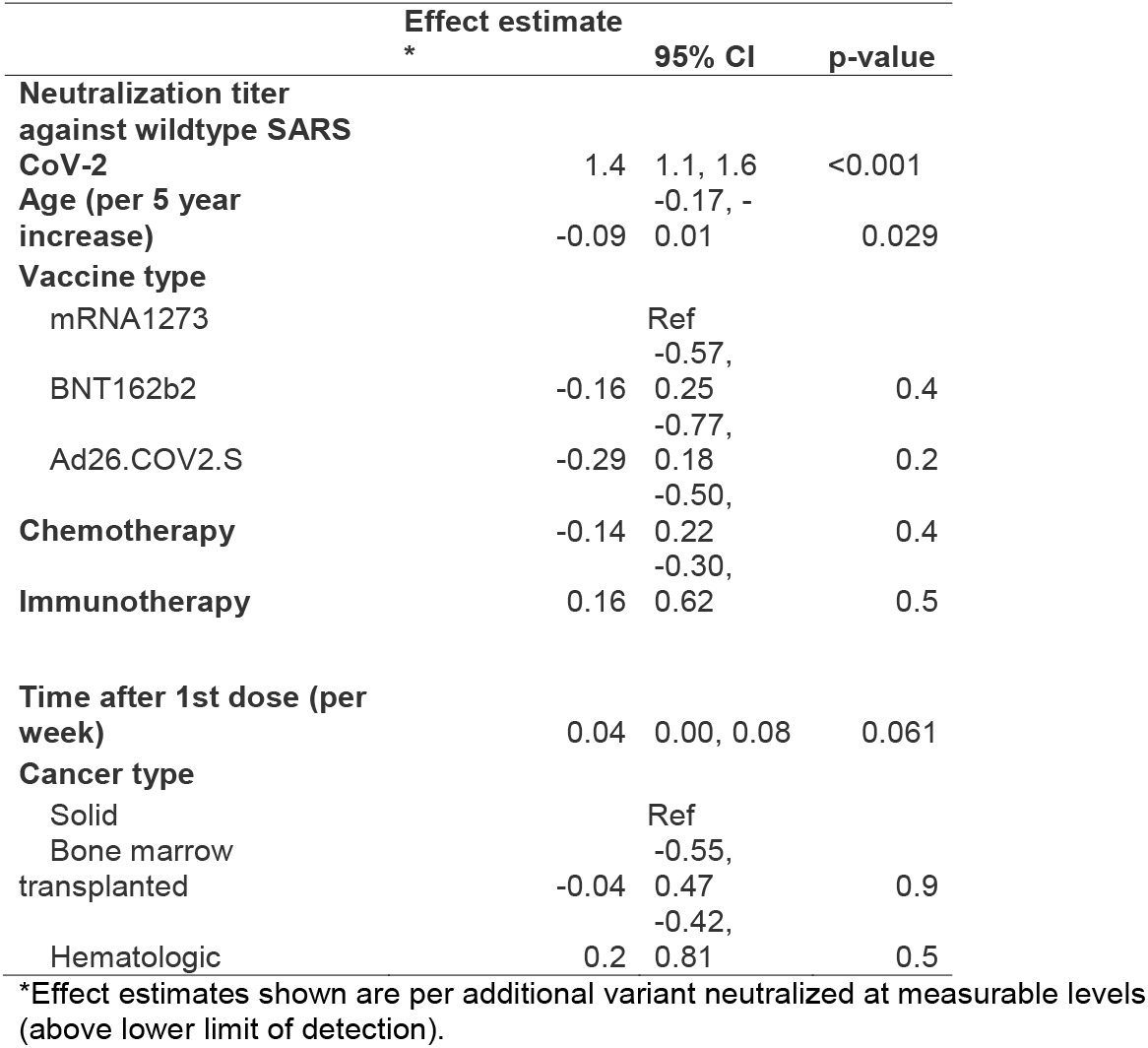
Multivariate regression analysis to identify correlates of breadth of neutralization.

### Anti-spike binding antibody concentrations are a surrogate for breadth

We measured total binding antibodies against SARS-CoV-2 spike (combined IgA/M/G) with a commercial (FDA EUA) assay and specific isotypes (IgA, IgG, IgM and combined IgA/M/G) of antibodies binding the receptor-binding domain with a validated assay we previously developed(Garcia-Beltran et al., 2021b; Wilfredo Garcia-Beltran et al., 2021). Anti-RBD responses were dominated by IgG isotype responses, and responses varied by vaccine as previously reported (Supplementary Figure 1). Wildtype neutralization was more robustly correlated with anti-RBD concentrations (Pearson R=0.63, 95% CI 0.53-0.71, p<0.001) than overall anti-spike responses (Pearson R=0.53, 95% CI 0.41-0.63, p<0.001) but both correlations were modest. A response comprised of more isotypes was not associated with greater breadth after adjusting for neutralization titer against wildtype virus as having a more isotype diverse response was associated with neutralization titer (p<0.001), as in other studies (Noval et al., 2021).

Anti-spike IgA/M/G total antibodies are easily measured in clinical practice, whereas neutralization is not. An anti-spike IgA/G/M titer >1000 U/ml on the Roche Elecsys FDA EUA assay was predictive of neutralization breadth (positive predictive value for neutralization of >2 variants at a titer >20 90%, negative predictive value 88%, overall sensitivity 95%, overall specificity 78%; Supplementary Figure 2).

### Additional ‘booster’ SARS-CoV-2 vaccines confer enhanced variant neutralization breadth

Since the magnitude of wildtype response associated with breadth, and booster doses increase the magnitude of wildtype response, we hypothesized that an additional homologous vaccine dose (or ‘booster’) elicits enhanced heterologous breadth. The safety of additional doses in this (Ref CANVAX) and prior studies (Ref RCT) has reported to be comparable to the primary series. In 13 participants with low baseline response (Supplementary Table 3), booster doses enhanced neutralization of alpha, beta, gamma and delta variants (Figure 2). Notwithstanding that these participants had low pre-booster titers (only 1 had measurable neutralization of any strain), the magnitude of neutralization of alpha, beta, gamma and delta variants was numerically higher than the overall cohorts who had received the full vaccine series.

**Figure 2:**
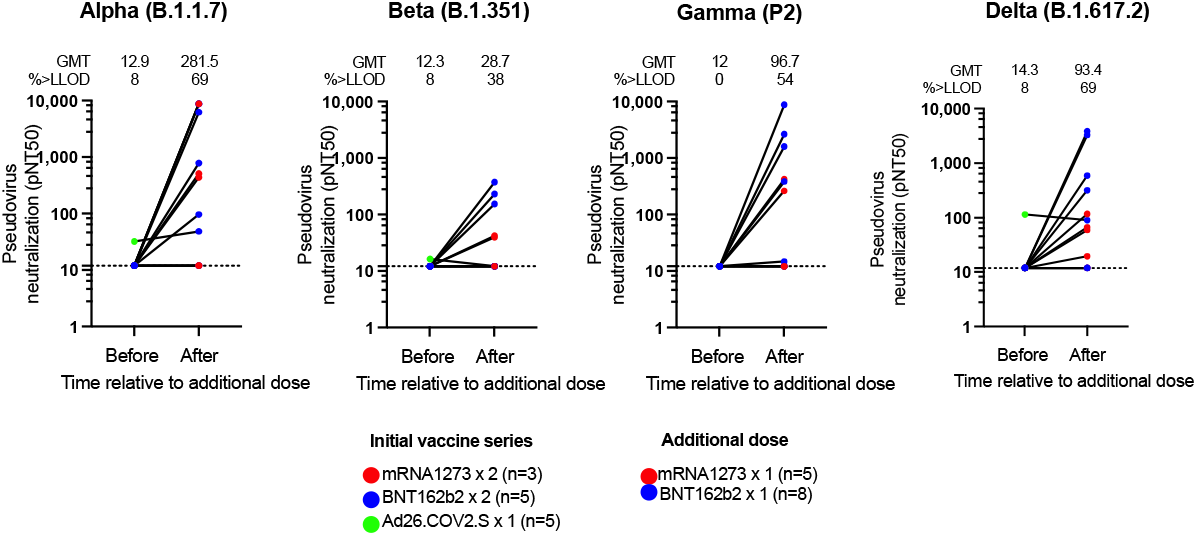
Effect of booster doses on neutralization of SARS CoV-2 viral variants in patients with cancer (n=13). The color of each dot indicates the initial vaccine series and additional vaccine as indicated in legend. The geometric mean titer (GMT) and proportion above the lower limit of detection (LLOD=12) is shown.

## Discussion

SARS-CoV-2 vaccination induces lower magnitude response in patients with cancer. Here we studied the breadth of response against SARS-CoV-2 variants as these represent the leading threat to vaccinated individuals. As in patients without cancer, vaccination with SARS-CoV-2 vaccines induces lower neutralization of variants, particularly beta, than wildtype. The vaccine types varied in magnitude of response but crucially, the magnitude of wildtype neutralization response was the primary correlate of breadth of neutralization. As predicted and concordant with another small study of four individuals (Iketani et al., 2021), booster doses even with wildtype vaccines increase breadth against viral variants.

These data have several potential clinical implications. First, to achieve the greatest neutralizing breadth, the most immunogenic vaccine may be preferred as the primary series for patients at high risk, where feasible. Second, differences in the effectiveness of the three FDA EUA vaccines in the USA (Naranbhai et al., 2021; Self, 2021) are likely accounted for by differences in magnitude of immunogenicity as opposed to differences in breadth per se, as each vaccine encodes largely conserved wildtype spike sequences. Finally, booster doses with wildtype vaccines would be expected to increase protection against variants in patients with cancer even as we await next generation of vaccines. This observation is likely because of boosting of polyclonal responses capable of binding conserved sites in current and predicted future variants. The magnitude of increase in variant naturalization appears to be robust, even when responses to initial vaccination are poor, suggesting a single booster dose may be adequate for most patients to overcome the apparent reduced priming of responses.

Whether these patients may require additional doses due to waning responses remains unclear.

There are several additional noteworthy observations. Increasing age was associated with reduced breadth of neutralization (independent of magnitude of response); concordant with impaired immune functions such as poorer somatic hypermutation, with age(Troutaud et al., 1999). Interestingly we observed a trend towards enhanced breadth over time, but this is likely small and likely not at the magnitude seen in individuals with natural infection who continue to accrue increased breadth over time potentially because of antigen persistence(Cho et al., 2021).

Interestingly, an anti-spike antibody titer higher than 1000 U/ml on the Roche Elecsys assay, an assay that is widely available in clinical practice, was a good surrogate for breadth. This may be a helpful threshold in counselling patients regarding need for boosters, notwithstanding limitations of inferring clinical protection from immunologic measures.

A key limitation of this study is the focus on in vitro immunogenicity.

Effectiveness of vaccines against variants is likely to be more complex involving differences in viral infectivity, exposure rates, transmissibility and possibly virulence in combination with largely humoral immune responses and cellular responses. This is illustrated by the exceptional success of the delta variant in transmission, but its relatively modest escape of neutralization. The number of individuals evaluated after booster doses is modest, but these represent the extra tail of the curve of patients who failed to make adequate responses after initial vaccination.

In conclusion, while current wildtype-based SARS CoV-2 vaccines induce lower magnitude responses in patients with cancer that show impaired neutralization of viral variants, boosting these responses can safely restore breadth against current viral variants.

## Data Availability

All data produced in the present study are available upon reasonable request to the authors

## Acknowledgments

We thank the Lambertus Family Foundation and Donald Glazer for support of the CANVAX study. We thank Andrea Nixon and the MGH Core laboratory for assistance in performing assays. We wish to thank Michael Farzan, PhD for providing ACE2-expressing 293T cells. A.J.I. and this study were supported by the Lambertus Family Foundation. A.B.B. was supported by the National Institutes for Drug Abuse (NIDA) Avenir New Innovator Award DP2DA040254, the MGH Transformative Scholars Program as well as funding from the Charles H. Hood Foundation.

## Author Contributions

Conceptualization, V.N., A.J.I.

Methodology, V.N., J.F.G., A.B.B, A.J.I., W.F.G.B.,

Formal Analysis V.N., K.J.S.D, E.C.L

Investigation V.N. K.J.S.D, E.C.L, O.O., W.F.G.B., C.B., A.B.B.

Data Curation V.N.

Writing-Original Draft V.N.

Writing-Review & Editing V.N., K.J.S.D, E.C.L, O.O., W.F.G.B., C.B., A.B., J.F.G., A.B.I., A.J.I.

Visualization V.N.

Supervision J.F.G., A.B.B, A.J.I.

Funding acquisition J.F.G., A.B.B, A.J.I.

## Inclusion and Diversity Statement

We worked to ensure gender balance in the recruitment of human subjects. We worked to ensure ethnic or other types of diversity in the recruitment of human subjects. One or more of the authors of this paper self-identifies as an underrepresented ethnic minority in science. One or more of the authors of this paper self-identifies as a member of the LGBTQ+ community. While citing references scientifically relevant for this work, we also actively worked to promote gender balance in our reference list.

## STAR Methods

### Patients

The CANVAX study enrolled consenting adult patients at the Massachusetts General Hospital Cancer Center between April 21 through July 21, 2021. Recruitment and enrolment procedures have been previously described. For this analysis we randomly selected participants without prior SARS CoV-2 infection (confirmed by anti-nucleocapsid antibody testing) who had received each of the three FDA EUA vaccines (by allocating random numbers to each participant and selecting the first 60).

Participants were samples >=14 days after their final dose of vaccine. This study was approved by the Mass General Brigham Human Research Committee (2021P000746).

### Neutralization assays

We used pseudovirus neutralization assay that we have previously described in detail elsewhere. In brief, pseudovirus neutralization titer 50 (pNT50) was calculated by taking the inverse of the serum concentration that achieved 50% neutralization of SARS-CoV-2 pseudotyped lentivirus particles entry into ACE2 expressing 293T cells. We introduced mutations corresponding to the SARS CoV-2 variants of concern shown in Supplementary Table 1 by site directed mutagenesis, and confirmed clones by sequencing.

### Binding antibody assays

We measured antibodies against the spike protein with the Roche Elecsys Anti-SARS-CoV-2 S assay (Roche Diagnostics, Indianapolis, USA), at the MGH Core Clinical laboratory, a CLIA lab. Anti-receptor binding domain antibodies were measured with an enzyme-linked immunosorbent assay (ELISA) that we have previously described.

### Statistical Methods

Analyses were performed in R (v4.05) using the *gtsummary* packages and lm() functions. We modelled log_10_ transformed pseudovirus neutralization titers as the dependent variable. We selected covariates found to associated with neutralization of wildtype SARS CoV-2 in the overall CANVAX study comprising >600 individuals these were: age, days post-vaccination, vaccine type, cancer type (categorized into solid, bone-marrow transplant or hematologic), receipt of chemotherapy in prior year and receipt of immunotherapy in prior year as the independent variables. Figures were rendered in GraphPad Prism v9.0.

## Supplemental Information

**Supplementary Figure 1:**
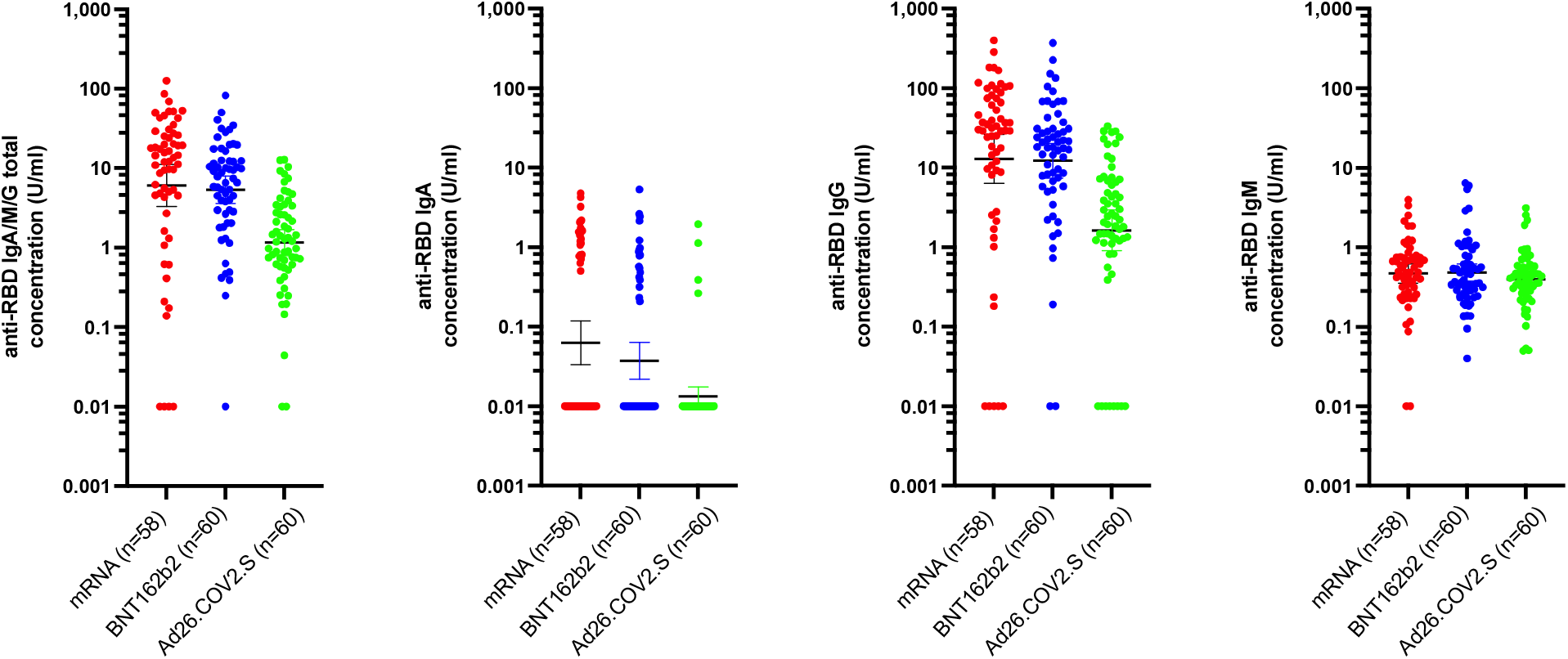
Concentration of antibodies against SARS-CoV-2 spike protein receptor binding domain (RBD) of a) IgA/M/G total, b) IgA, c) IgG and d) IgM isotypes.

**Supplementary Figure 2:**
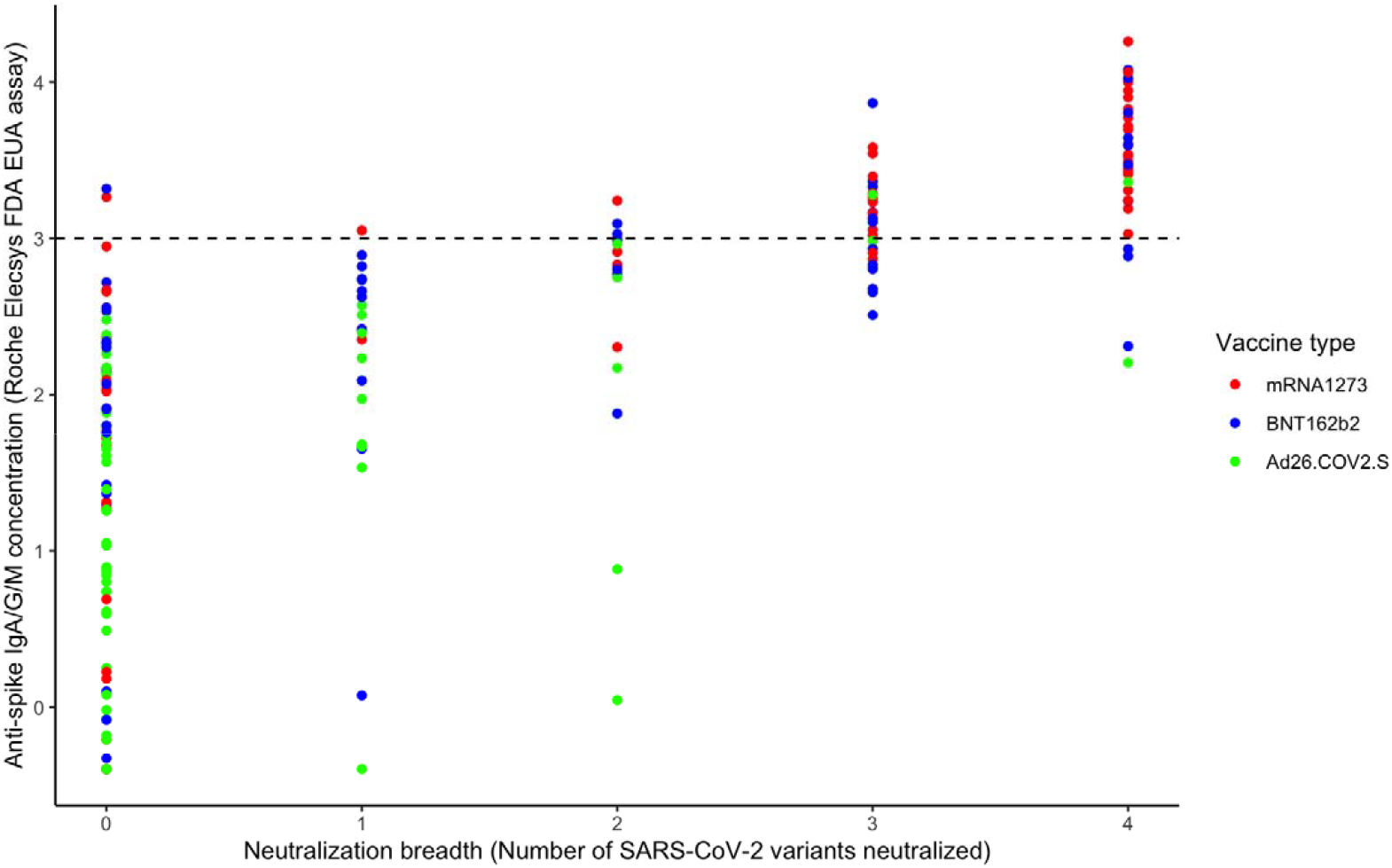
Association between anti-spike IgA/G/M concentration (Roche Elecsys assay) and neutralization breadth. Variant neutralization was defined as neutralization >20 for each variant. A horizontal line is drawn at an antibody concentration of 3log_10_ (1000 U/mL), which corresponds to a positive predictive value(PPV) of 90% for >2 variants neutralized.

**Supplementary Table 1:**
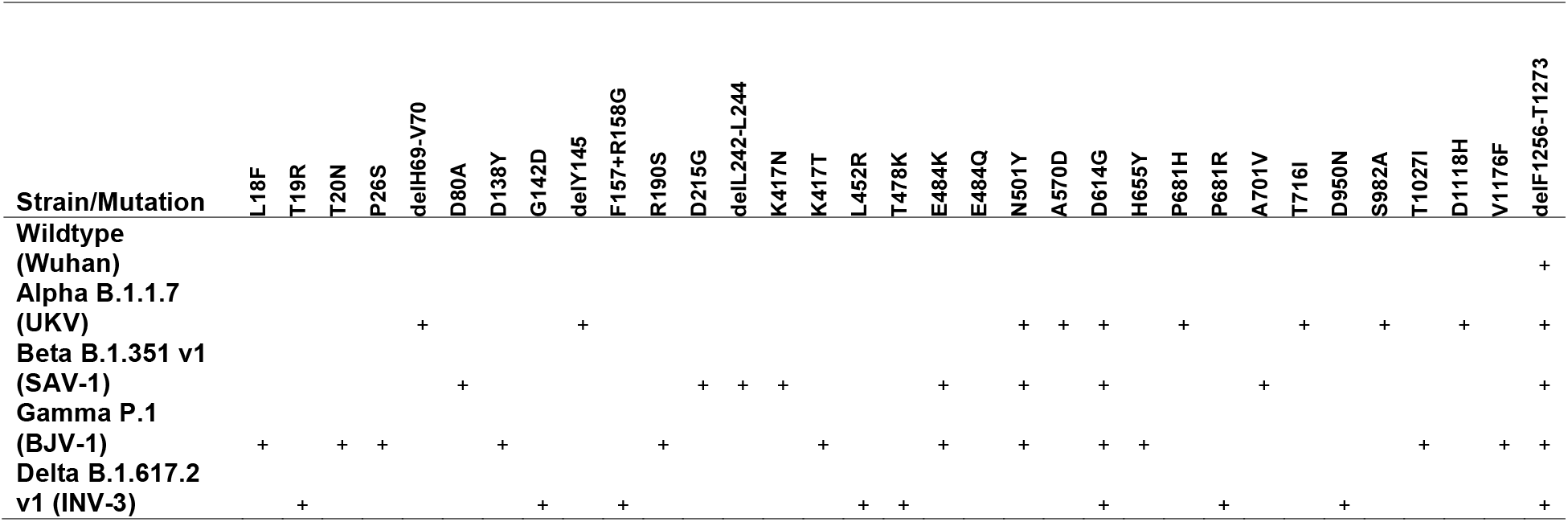
Summary of mutations in SARS CoV-2 strains used in this study.

**Supplementary Table 2:**
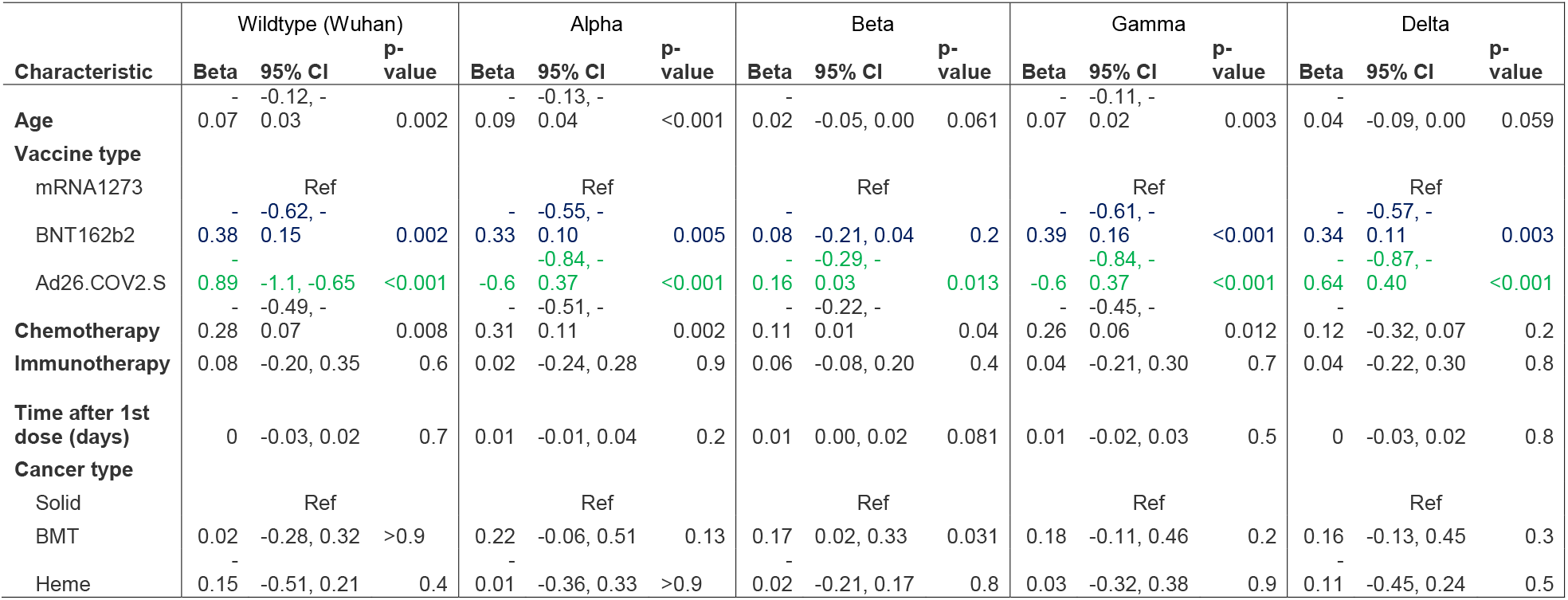
Correlates of neutralization of viral variants in patients with cancer. Shown are results of multivariate regression model including the covariates shown, with log10 pseudovirus neutralization of each viral variant as the response variable.

**Supplementary Table 3:** Characteristics of patients who received booster vaccine doses.

|  | **Cancer type, therapy** | **Initial Vaccine series** | **Additional vaccine dose type** | **Interval between initial and additional dose (days)** |
| --- | --- | --- | --- | --- |
| 1 | Cutaneous Squamous Cell Carcinoma on immunotherapy | Ad26.COV2.S | BTN162b2 | 50 |
| 2 | Breast cancer previously on chemotherapy | Ad26.COV2.S | BTN162b2 | 81 |
| 3 | Breast cancer and marginal zone lymphoma on lenalidomide (no B-cell depleting agent) | Ad26.COV2.S | BTN162b2 | 81 |
| 4 | Oral squamous cell cancer receiving targeted therapy and steroids | Ad26.COV2.S | BTN162b2 | 104 |
| 5 | Lung cancer on immunotherapy and steroids | Ad26.COV2.S | mRNA1273 | 86 |
| 6 | Breast cancer on active chemotherapy | mRNA1273 | BTN162b2 | 96 |
| 7 | Bone marrow transplanted on immunosuppression | mRNA1273 | mRNA1273 | 79 |
| 8 | Lung cancer, on chemotherapy and steroids | mRNA1273 | mRNA1273 | 115 |
| 9 | oral squamous cell cancer receiving chemotherapy, immunotherapy and steroids | BTN162b2 | BTN162b2 | 73 |
| 10 | lung cancer (early stage) receiving chemoradiation | BTN162b2 | BTN162b2 | 80 |
| 11 | Breast cancer on targeted therapy | BTN162b2 | BTN162b2 | 108 |
| 12 | Oral squamous cell cancer receiving chemoradiation | BTN162b2 | mRNA1273 | 78 |
| 13 | Pancreatic cancer receiving chemotherapy | BTN162b2 | mRNA1273 | 88 |

